# Fear avoidance beliefs limit lumbar spine flexion during object lifting in pain-free adults

**DOI:** 10.1101/2020.04.01.20049999

**Authors:** D Knechtle, S Schmid, M Suter, F Riner, G Moschini, M Senteler, P Schweinhardt, ML Meier

## Abstract

There is a long-held belief that physical activities such as lifting with a flexed spine is generally harmful for the back and can cause low back pain (LBP), potentially nurturing fear avoidance beliefs underlying pain-related fear. In chronic LBP patients, pain-related fear has been shown to be associated with reduced lumbar range of motion during lifting, indicating distinct and probably protective psychomotor responses to pain. However, despite short term beneficial effects for tissue health, recent evidence suggests that maintaining a protective trunk movement strategy may also pose a risk for (persistent) LBP due to possible pro-nociceptive consequences of altered spinal kinematics, reflected by increased loading on lumbar tissues and persistent muscle tension. Yet, it is unknown if similar psychomotor interactions already exist in pain-free individuals which would yield potential insights into how a person might react when they experience LBP. Therefore, the aim of this study is to test the impact of pain-related fear on spinal kinematics in a healthy cohort of pain-free adults without a history of chronic pain. The study subjects (N=57) filled out several pain-related fear questionnaires and were asked to perform a lifting task (5kg-box). High-resolution spinal kinematics were assessed using an optical motion capturing system. Time-sensitive analyses were performed based on statistical parametric mapping. The results demonstrated time-specific and negative relationships between self-report measures of pain-related fear and lumbar spine flexion angles during lifting, yielding important implications regarding unfavorable psychomotor interactions that might become relevant in a future LBP incident.

## 1. Introduction

Emotions and beliefs shape how humans move and vice versa [26,38]. A prime example for this interplay is pain: people move differently in (the expectation of) pain, and conversely, dysfunctional or degraded movement can enhance pain [9,21]. This particularly applies to body parts thought to require superior protection such as the back [11,12,20]. Common beliefs are that the back is easily injured and that the healing process is long [11]. Such beliefs can increase protective behaviors, including control of posture and avoidance of daily activities, potentially aggravating disability and pain in the long term [12,59,63].

Activities that are believed by many to be harmful for the back, and even a potential cause of low back pain (LBP), include lifting with a flexed spine [6,17,50]. However, recent studies have not found convincing evidence that the spine should not be flexed during lifting to prevent LBP [13,29,50,60,62,64]. On the contrary, maintaining a protective strategy, e.g. by keeping a neutral spine (through “stiffening” the lumbar spine) during lifting, has been shown to be associated with increased muscle co-contraction and mechanical loading on spinal tissues, which might result in ongoing nociceptive input in the long term [8,16,19,32,34,39,47,59]. Yet, many health care professionals still promote lifting with a neutral spine as the safer lifting technique [42,50,62], potentially nurturing erroneous beliefs underlying pain-related fear (e.g. that the back is in danger when flexed). In support of this notion recent evidence indicates an implicit bias towards “lifting with a flexed spine is dangerous”, compared to lifting with a neutral spine, in patients with persistent LBP as well as in the pain-free individuals [5,6]. Brain research further supports this by demonstrating distinct relationships between self-reports of pain-related fear and fear-related neural activity during observation of daily activities such as lifting with a flexed spine in LBP and pain-free subjects [35,36,57]. However, the underlying psychomotor interactions between pain-related fear and spinal motion are largely unknown and need to be elucidated to disentangle possible clinically relevant relationships between pain-related fear, spinal motion and negative outcomes such as persistent LBP and disability. With respect to this, there is a lack of studies measuring lumbar spine flexion during lifting mimicking real life settings [50], especially with regards to psychomotor interactions in people with and without LBP. First insights came from a cross-sectional study demonstrating that elevated pain-related fear of lifting is linked to reduced sagittal plane lumbar range of motion (ROM) in chronic non-specific LBP patients during a lifting task [34]. The results of this study indicate an unfavorable “trunk stiffening” strategy in fearful LBP patients [39,59]. However, based on the reportedly pre-existing fear avoidance beliefs in pain-free individuals [5,36], it would be crucial to know whether pain-related fear also affects spinal motion in pain-free subjects, yielding potential insights into how a person might react when they experience LBP.

Therefore, using high-resolution spinal kinematics, we investigated whether pain-related fear affects lumbar motion during lifting in pain-free adults. In addition to conventional ROM analyses, we applied statistical parametric mapping to obtain time-sensitive information regarding changes of spinal motion [43].

## 2. Methods

### 2.1. Participants

Sixty-one pain-free and healthy adults (males/females: 31/30; age: 29.5 ± 6.9 years) were enrolled in this study. Recruitment took place between January and November 2019, using the following inclusion criteria: age between 18-60 years, no acute or recurrent LBP within the past 3 months, no history of chronic pain, no prior spine surgery, no history of psychiatric or neurological disorders, not being pregnant, no consumption of alcohol or drugs within the past 24 hours and a BMI of lower or equal to 30 kg/m^2^. The study protocol was approved by the local ethics committee (Kantonale Ethikkommission Zürich, EK-01/2019/PB_2018_01001) and conformed to the Declaration of Helsinki. All participants provided written informed consent prior to any study-related activities. They were invited for a single visit at the local university hospital, where they completed several questionnaires and underwent a three-dimensional optical full-body movement analysis.

### 2.2. Questionnaires

Participants completed the two following questionnaires assessing pain-related fear:

1. The modified 17-item German version of the Tampa Scale for Kinesiophobia (TSK) for the general population (TSK-G) assesses subjective ratings of pain-related fear of movement/(re)injury due to physical activity and kinesiophobia using a 4-point Likert scale ranging from 1 = “strongly disagree” to 4 = “strongly agree” [22]. It includes questions such as “If I had pain, I would feel better if I was physically active” and therefore measures more general aspects of pain-related fear. Psychometric research indicated a sufficient reliability (Cronbach’s *α* = 0.78); the score range lies between 17 (low level of kinesiophobia) and 68 (high level of kinesiophobia) [22].
2. The Photograph Series of Daily Activities - Short electronic Version (PHODA-SeV) is a tool for measuring the perceived harmfulness of certain movements. Pictures of different daily tasks are presented to the participants who are then asked to imagine themselves in the shown situations and indicate how harmful they think these activities would be to their back on a scale from 0 to 100 (0 = not harmful at all; 100 = extremely harmful). The internal consistency of the total score on the PHODA-SeV, as indicated by Cronbach’s α, was reported as 0.98 and the corrected item-total correlations ranged between 0.42 and 0.82, indicating that each item was moderately to highly related to the other items [46]. For the current study, we chose a priori the overall score (PHODA-total) and the score of the item showing a person lifting a flowerpot with a bent back (PHODA-lift) as variates of interest. Lifting a flowerpot best reflects a typical lifting task and has demonstrated a specific relationship between harmfulness ratings and the lumbar lifting ROM in chronic LBP patients [34].

To investigate potential differences or shared variance between self-reports of pain-related fear and general anxiety, we used the State-Trait Anxiety Inventory (STAI), which includes two subscales [54]. The State Anxiety Scale (S-Anxiety) assesses current levels of anxiety, whereas the Trait Anxiety Scale (T-Anxiety) evaluates more stable aspects of anxiety such as “anxiety proneness” [27].

### 2.3. Full-body movement analysis

Participants were equipped with 58 retro-reflective skin markers placed by a physiotherapist or movement scientist with experience in palpation according to a previously described marker configuration [51]. To enable detailed tracking of spinal motion, this configuration included markers placed on the spinous processes of C7, T3, T5, T7, T9, T11, L1 to L5 and S1 (Figure 1).

**Figure 1.**
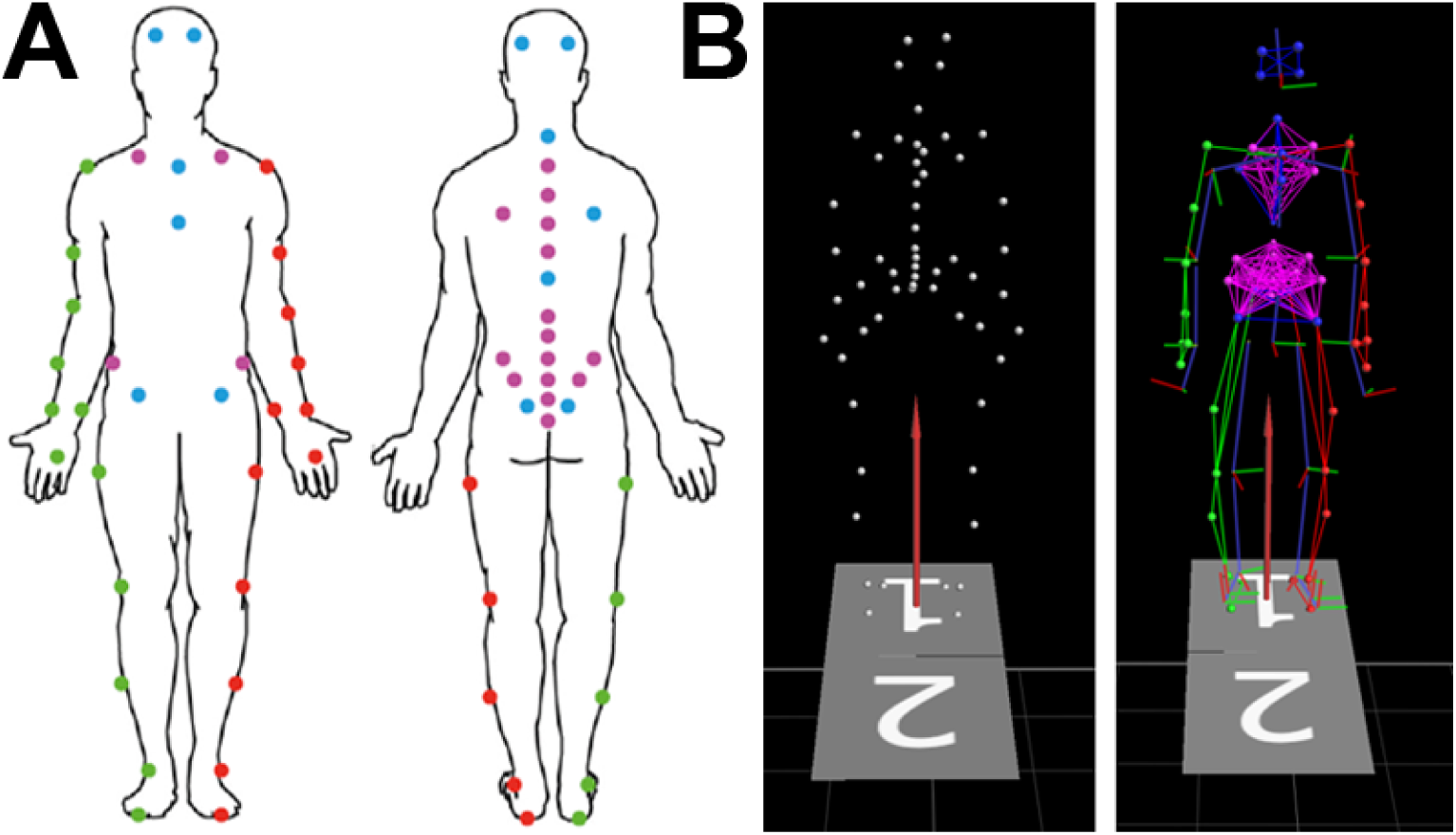
A = Full body marker placement according to Schmid et al. [51] including head, pelvis, thorax, spine, shoulder, elbow, wrist, arms and lower extremities. Markers placed on the spinous processes of C7, T3, T5, T7, T9, T11, L1 to L5 and S1 were used for tracking of spinal motion. B = Vicon interface showing the captured and reconstructed 3D marker positions before (left) and after labeling and Plug-in Gait model calculations (right).

Participants were then asked to perform a series of activities of daily-living including upright standing and sitting on a chair, bending forward and backward from an upright standing position without bending their knees, standing up from a chair and sitting down on a chair with free hanging arms, lifting-up and putting-down a 5 kg-box that was placed 15 cm in front of the subjects’ feet, walking and running on a level ground as well as climbing up and down a stair with four steps. No further instructions were given to ensure individual and natural movements at self-selected speeds. Apart from standing, sitting and bending (performed once), all activities were repeated until five valid trials were collected. For familiarization with the tasks the participants practiced the activities prior to the actual testing. Testing was repeated if the participants violated the task instructions, resulting in non-valid trials. For the current study only data from bending and lifting activities were considered.

Three-dimensional marker positions were tracked using a 20-camera optical motion capturing system (Vicon UK, Oxford, UK) at a sampling frequency of 200 Hz.

### 2.4. Data reduction and outcome parameters

Motion capture data were pre-processed using the software Nexus (version 2.8.1, Vicon UK, Oxford, UK), involving marker reconstruction and labeling, gap filling and filtering of the marker trajectories as well as setting of temporal events for the identification of the relevant data sections.

Post-processing was carried out with a custom-built MATLAB routine (R2019a, MathWorks Inc., Natrick, MA, USA). In a first step, marker data were cropped according to the temporal events set during pre-processing or defined using a previously described event-detection algorithm (i.e. end point of the lifting-up as well as starting point of the putting-down activities) [56].

Lumbar angles of the bending forward activity as well as lumbar and thoracic angles of the lifting activity were calculated based on the trajectories of the L1 to S1 and C7 to T11 markers, respectively, using a combination of a quadratic polynomial and a circle fit function [52]. For the lifting activity, we additionally applied a quintic polynomial function to all sagittal plane spinal marker trajectories (i.e. C7 to S1) to derive segmental angles (angles between the normal lines passing through the individual marker positions) of the L1/L2, L2/L3, L3/L4, L4/L5 and L5/S1 spinal units [23,24]. Information on maker placement accuracy and soft tissue artifacts can be found elsewhere [52,67]. For time-sensitive analyses, continuous angles from the lifting activity were time-normalized on 101 points (time window: 0 – 100%) and averaged across all five trials (per subject). To obtain ROM values for the analyzed tasks, continuous angles were reduced to a discrete flexion ROM value (averaged across the five trials), i.e. angle difference between upright standing and maximal deviation from the starting position. All angles were expressed in degrees (°).

The continuous lumbar lordosis angles in the sagittal plane during lifting-up and putting-down a box were the primary outcomes. Secondary outcomes included the continuous lumbar segmental angles and thoracic kyphosis angles in the sagittal plane during lifting-up and putting-down a box.

### 2.5. Statistical analysis

Statistical calculations were performed using SPSS (version 23, SPSS Inc., Chicago, IL, USA) and the Python-based software package for one-dimensional Statistical Parametric Mapping (SPM: spm1d-package, www.spm1d.org) [44]. Prior to any inferential analyses, data were tested for normality using the D’Agostino’s K2 test (SPM function *spm1d*.*stats*.*normality*.*k2*.*ttest*) for the continuous spinal angles and the Shapiro-Wilk test and Q-Q plot inspection for measures of pain-related fear. In case of non-normal distribution of the questionnaire data, Spearman’s rank correlation coefficient was used for correlation analysis. To investigate potential relationships between continuous spinal angles and measures of pain-related fear, we conducted multiple linear regression analyses (SPM function *spm1d*.*stats*.*glm*) using measures of pain-related fear as regressors of interest and age, gender and bending ROM as nuisance variables (as they have been shown to possibly influence lumbar and thoracic curvature angles [2,25,33]). For each measure of pain-related fear, a separate regression analysis for the lifting-up and putting-down phases was performed and the output statistic SPM{t} was calculated at each of the 101 time points.

Tests were based on the null hypothesis, i.e. there are no relationships between continuous spinal angles and the respective measure of pain-related fear. Assuming principles of Random Field Theory that were validated for 1D data [45,46], statistical significance was determined by a critical SPM{t}-threshold at which only α% (5%) of smooth random curves would be expected to traverse [43]. This leads to “supra-threshold clusters” that characterize significant time-specific positive or negative relationships between spinal angles and pain-related fear. For a better interpretability of the effect sizes, the respective t-statistics were transformed to correlation coefficients (r) based on the following formula:

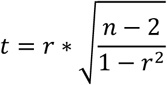

Multiple comparisons correction was performed for primary outcomes and was based on a false discovery rate (FDR) of 5% [3] (including six separate tests for TSK-G, PHODA-total, PHODA-lift regressors and continuous lumbar lordosis angles in lifting-up and putting-down phases).

To compare the actual data in pain-free adults with similar analyses recently performed in chronic LBP patients [34], we conducted the following complementary analyses: (1) multiple regression analyses including the TSK-G score (as a measure of general pain-related fear) as nuisance variable (in addition to age, gender and bending ROM) to test if movement-specific pain-related fear (PHODA items) explains additional variance in spinal kinematics during lifting after accounting for linear effects of the TSK-G score (section 3.6) and (2) correlation analyses between the lumbar ROM during lifting and measures of pain-related fear (TSK-G and PHODA items, section 3.7)

## 3. Results

### 3.1. Recruitment and subject characteristics

Four subjects had to be excluded from the analysis, resulting in a final sample of 57 pain-free healthy adults (males/females: 30/27; age: 29.5 ± 7.0 years; mass: 67.9 ± 11.8 kg; height: 174.4 ± 8.9 cm; BMI: 22.2 ± 2.6 kg/m^2^). The reasons for the exclusions were technical issues that led to the loss of the kinematic data (1 subject), conceptual misunderstanding of the PHODA questionnaire (1 subject, stating having switched the endpoints of the scale) and a hyperlordosis of the lumbar spine in neutral position with an angle of > 68° [14,28] (2 subjects).

### 3.2. Questionnaire data

The analysis of the PHODA harmfulness ratings indicated similar threat values for the a priori chosen item PHODA-lift and the items “shoveling soil” (PHODA-shoveling) and “falling backwards” (PHODA-falling) (see Table 4). We therefore added the latter two items post-hoc in the correlation analysis and performed exploratory time-sensitive regression analyses (see section 3.5).

**Table 4.** Spearman’s rank correlations (r) between scores on individual PHODA-SeV items and lumbar range of motion during lifting, sorted according to the mean threat value in descending order. Reported are mean ± SD and median with interquartile range (IQR).

| ID | Description | Score on item |  | Lifting ROM |  |
| --- | --- | --- | --- | --- | --- |
|  |  | Mean (SD) | Median (IQR) | Correlation | p-value |
| 1 | Shoveling soil | 51.3 (29.5) | 50 (29-80) | -0.121 | 0.370 |
| 38 | Falling backwards | 49.5 (25.3) | 53 (33-69) | -0.380 | **0.004** |
| 3 | Lifting pot, bent back | 47.5 (28.5) | 50 (24-73) | -0.113 | 0.404 |
| 10 | Lifting beer crate, bent back | 31.7 (22.9) | 30 (12-49) | -0.143 | 0.287 |
| 21 | Taking box from shelf above head | 30.2 (27.1) | 24 (7-52) | -0.080 | 0.555 |
| 31 | Lifting toddler | 29.9 (20.8) | 29 (11-47) | -0.124 | 0.357 |
| 11 | Carrying bag, one hand | 28.1 (21.6) | 27 (11-44) | -0.055 | 0.684 |
| 40 | Drilling hole above head | 27.6 (20.7) | 27 (10-41) | -0.008 | 0.953 |
| 32 | Carrying child on hip | 27.5 (20.1) | 25 (12-43) | 0.007 | 0.958 |
| 16 | Vacuum cleaning | 26.9 (23.4) | 20 (7-42) | -0.124 | 0.358 |
| 13 | Carrying rubbish, one hand | 24.0 (20.2) | 23 (6-35) | -0.013 | 0.925 |
| 4 | Picking up, bent | 22.4 (23.9) | 13 (3-35) | -0.062 | 0.644 |
| 39 | Mowing lawn | 22.3 (18.3) | 22 (3-36) | 0.083 | 0.540 |
| 17 | Mopping floor | 18.6 (16.1) | 15 (7-26) | -0.014 | 0.915 |
| 20 | Back bending | 18.2 (20.5) | 10 (3-29) | 0.162 | 0.229 |
| 22 | Trampoline jumping | 17.9 (19.7) | 10 (3-30) | 0.005 | 0.968 |
| 25 | Making bed | 17.5 (18.5) | 11 (4-27) | -0.001 | 0.993 |
| 9 | Lifting basket, stairs | 17.4 (16.1) | 12 (3-29) | 0.043 | 0.750 |
| 29 | Cleaning windows above head | 16.7 (16.9) | 11 (5-25) | -0.028 | 0.834 |
| 23 | Rope skipping | 16.1 (16.7) | 11 (3-28) | 0.037 | 0.786 |
| 19 | Back twisting | 15.8 (15.5) | 12 (2-25) | 0.223 | 0.095 |
| 34 | Running | 15.6 (15.0) | 10 (3-29) | -0.001 | 0.995 |
| 12 | Carrying bag, both hands | 15.3 (13.1) | 14 (5-24) | 0.023 | 0.864 |
| 18 | Leg stretching | 15.3 (15.1) | 11 (3-25) | 0.084 | 0.533 |
| 24 | Abdominal exercises | 15.0 (17.1) | 8 (3-23) | 0.116 | 0.391 |
| 33 | Doing dishes | 13.5 (14.6) | 8 (2-23) | 0.087 | 0.522 |
| 7 | Ironing, standing | 13.2 (19.2) | 5 (0-19) | 0.100 | 0.461 |
| 14 | Clear dishwasher | 12.6 (13.5) | 8 (3-20) | -0.059 | 0.665 |
| 37 | Cycling, looking aside | 12.5 (16.8) | 7 (2-18) | -0.034 | 0.804 |
| 2 | Lifting pot, squat | 11.8 (10.8) | 10 (0-20) | -0.111 | 0.413 |
| 36 | Cycling from kerb | 11.5 (16.1) | 7 (1-15) | -0.011 | 0.938 |
| 15 | Taking from cupboard | 11.3 (14.1) | 7 (0-16) | 0.147 | 0.276 |
| 5 | Picking up, squat | 11.0 (15.4) | 4 (0-17) | 0.029 | 0.828 |
| 30 | Riding bike, bumpy | 10.9 (17.2) | 6 (0-12) | 0.063 | 0.641 |
| 6 | Taking box, twisted back | 10.9 (14.3) | 5 (0-16) | 0.096 | 0.477 |
| 26 | Getting out of bed | 8.8 (10.6) | 4 (1-14) | 0.126 | 0.350 |
| 8 | Ironing, sitting | 7.2 (9.7) | 3 (0-11) | 0.105 | 0.435 |
| 28 | Walking down stairs | 6.7 (9.4) | 3 (0-7) | 0.051 | 0.708 |
| 35 | Walking | 5.2 (7.5) | 2 (0-10) | 0.033 | 0.810 |
| 27 | Walking up stairs | 3.9 (6.2) | 1 (0-5) | 0.004 | 0.976 |
|  | Phoda-total | 19.2 (12.2) | 19.1 (8-28) | -0.027 | 0.845 |

Q-Q plots inspection and the Shapiro-Wilk test indicated non-normality for the PHODA-lift (p = 0.019) and PHODA-shoveling (p = 0.022) as well as for the T-Anxiety (p = 0.002) and S-Anxiety (p = 0.001) score distributions. The PHODA-total, PHODA-falling and TSK scores were normally distributed (p > 0.05). Mean scores were 31.8 (SD=±5.5) for the TSK-G, 37.5 (SD=±6.5) for the T-Anxiety and 30.4 (SD=±7.5) for S-Anxiety. Mean values for each PHODA item are listed in Table 4. The T-Anxiety score moderately correlated with the PHODA-falling (Spearman’s r = 0.244, p = 0.034) and TSK-G (r = 0.233, p = 0.040) scores. No significant correlations were found between the TSK-G and PHODA-total, PHODA-lift, PHODA-shoveling and PHODA-falling scores (r’s < 0.16, p’s > 0.13). The results of the correlation analyses are summarized in Table 1.

**Table 1.**
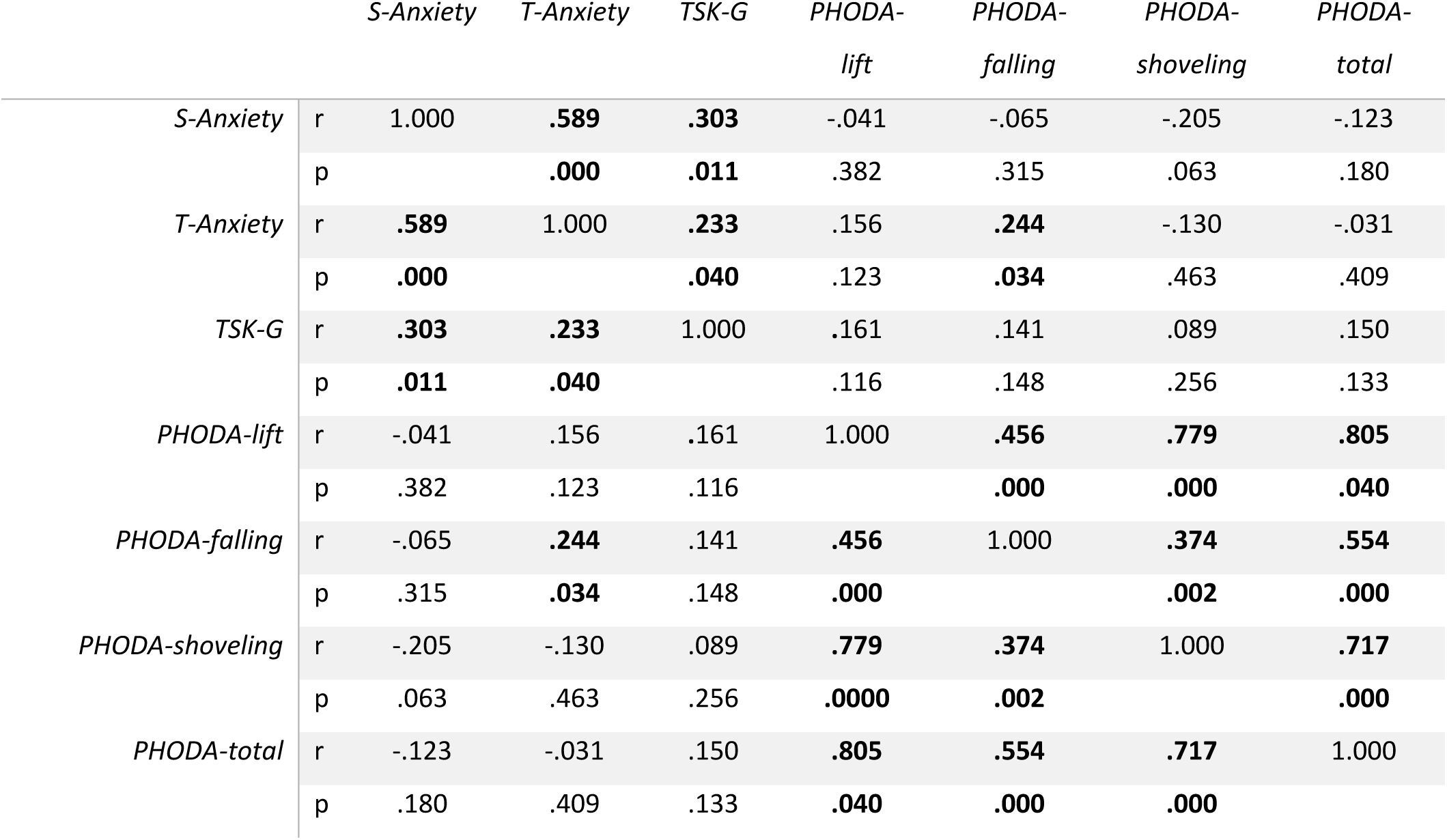
Spearman’s rank correlations (r) between the different questionnaires and PHODA items. Tampa Scale of Kinesiophobia for the general population (TSK-G). State and Trait Anxiety Inventory (S-Anxiety. T-Anxiety). PHODA items: lifting a flowerpot (PHODA-lift). falling backwards on the grass (PHODA-falling). shoveling soil (PHODA-shoveling). p < 0.05 (bold).

|  |  | *S-Anxiety* | *T-Anxiety* | *TSK-G* | *PHODA-lift* | *PHODA-falling* | *PHODA-shoveling* | *PHODA-total* |
| --- | --- | --- | --- | --- | --- | --- | --- | --- |
| *S-Anxiety* | r | 1.000 | **.589** | **.303** | -.041 | -.065 | -.205 | -.123 |
|  | p |  | **.000** | **.011** | .382 | .315 | .063 | .180 |
| *T-Anxiety* | r | **.589** | 1.000 | **.233** | .156 | **.244** | -.130 | -.031 |
|  | p | **.000** |  | **.040** | .123 | **.034** | .463 | .409 |
| *TSK-G* | r | **.303** | **.233** | 1.000 | .161 | .141 | .089 | .150 |
|  | p | **.011** | **.040** |  | .116 | .148 | .256 | .133 |
| *PHODA-lift* | r | -.041 | .156 | .161 | 1.000 | **.456** | **.779** | **.805** |
|  | p | .382 | .123 | .116 |  | **.000** | **.000** | **.040** |
| *PHODA-falling* | r | -.065 | **.244** | .141 | **.456** | 1.000 | **.374** | **.554** |
|  | p | .315 | **.034** | .148 | **.000** |  | **.002** | **.000** |
| *PHODA-shoveling* | r | -.205 | -.130 | .089 | **.779** | **.374** | 1.000 | **.717** |
|  | p | .063 | .463 | .256 | **.0000** | **.002** |  | **.000** |
| *PHODA-total* | r | -.123 | -.031 | .150 | **.805** | **.554** | **.717** | 1.000 |
|  | p | .180 | .409 | .133 | **.040** | **.000** | **.000** |  |

### 3.3. Relationships between TSK-G, PHODA-lift, PHODA-total and continuous lumbar and thoracic angles during lifting

Multiple linear regression analysis revealed a statistically significant negative relationship between the PHODA-lift score and continuous lumbar angles during the lifting-up (time window: 9-92%, -0.313 ≤ r ≥ - 0.310, p_FDR_ = 0.007) and putting-down (time window: 17-60%, -0.315 ≤ r ≥ -0.306, p_FDR_ = 0.028) phases (Figure 2A and 2B, Table 2). No relationships were found for TSK-G, PHODA-total and continuous lumbar angles nor for any of the three scores and continuous thoracic angles (p_FDR_ > 0.05).

**Table 2.**
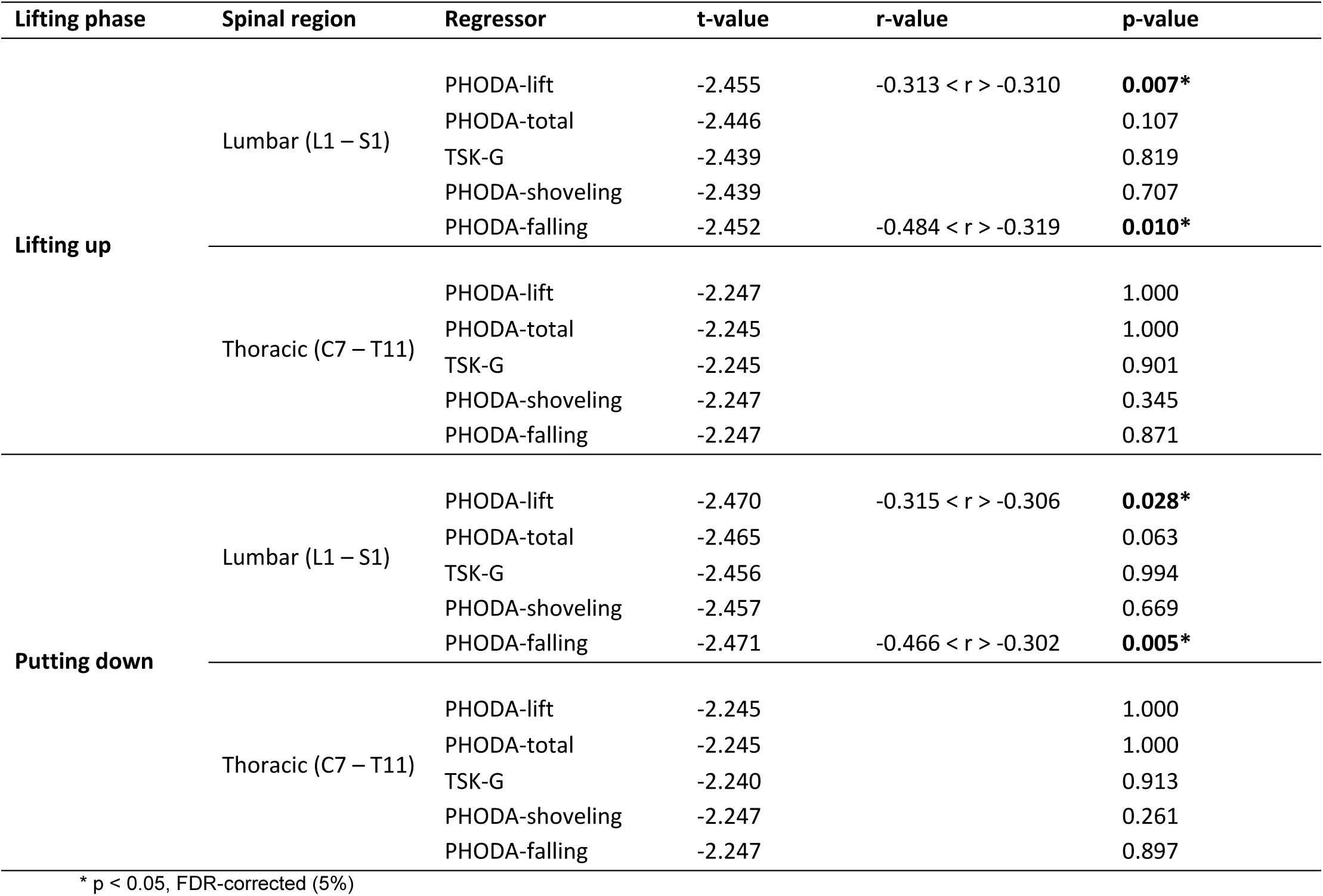
Relationships between measures of pain-related fear and continuous lumbar and thoracic angles during lifting

**Figure 2.**
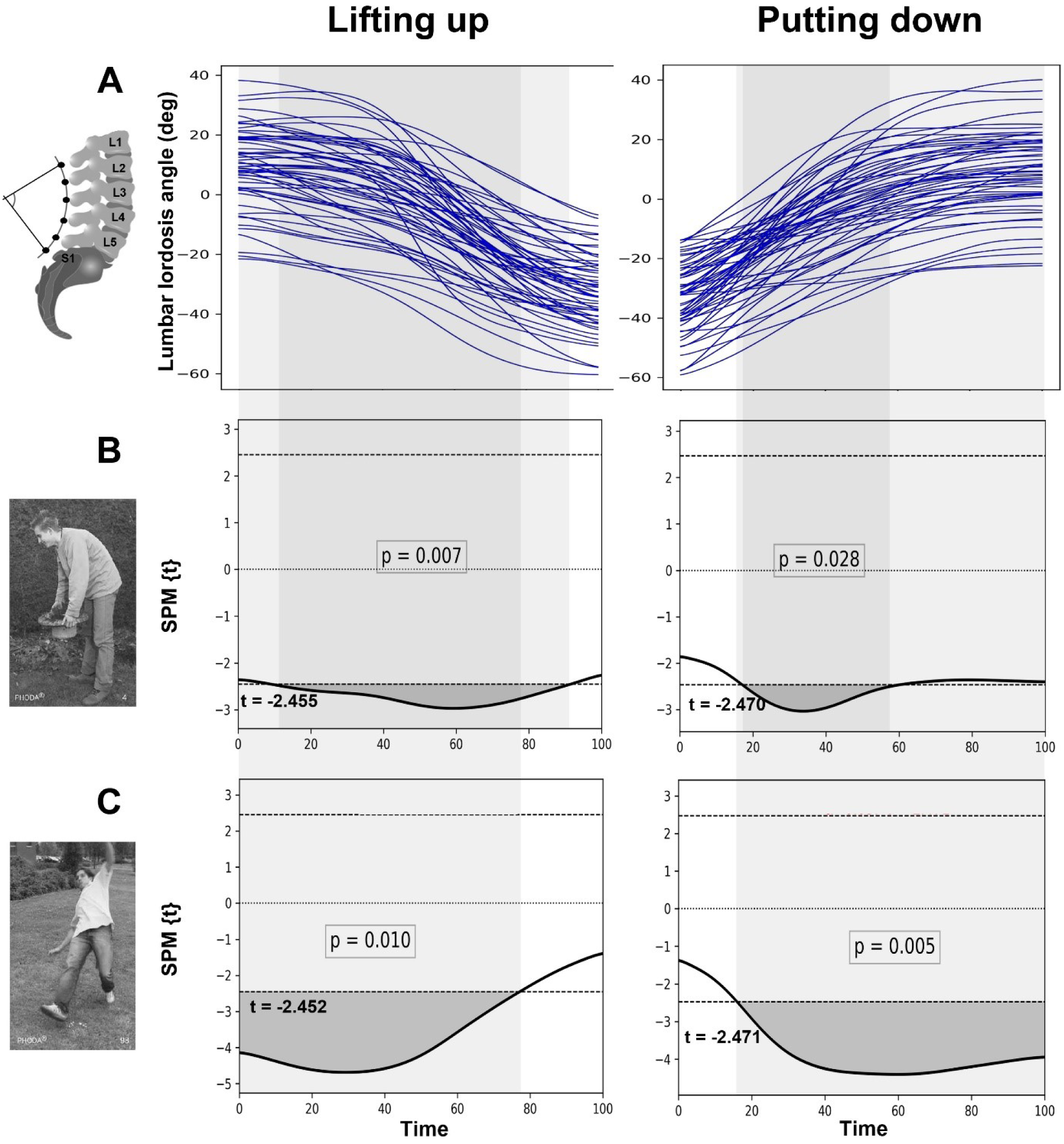
A = Individual (N = 57) continuous lumbar angles during lifting-up (left) and putting-down (right) phases. X-axis: time normalized on 101 points (time window: 0 – 100%). B, C = t-statistics with supra-threshold clusters reflecting significant time-specific negative relationships between lumbar angles during the two phases and the PHODA-lift (B) and PHODA-falling (C) score, revealed by SPM1D multiple linear regression.

### 3.4. Relationships between PHODA-lift and continuous lumbar segmental angles during lifting

Multiple regression analyses with the continuous angles of the lumbar segments L1/L2, L2/L3, L3/L4, L4/L5 and L5/S1 as dependent variables revealed that the time-specific relationships between the lumbar angle and the PHODA-lift score were driven by motion in the L4/L5 segment during the lifting-up (time window: 0-61%, -0.333 ≤ r ≥ -0.315, p_uncorr_ = 0.021) as well as the putting-down (time window: 29-100%, 0.354 ≤ r ≥ -0.305, p_uncorr_ = 0.012) phases (Figure 3A and 3B, Table 2).

**Figure 3.**
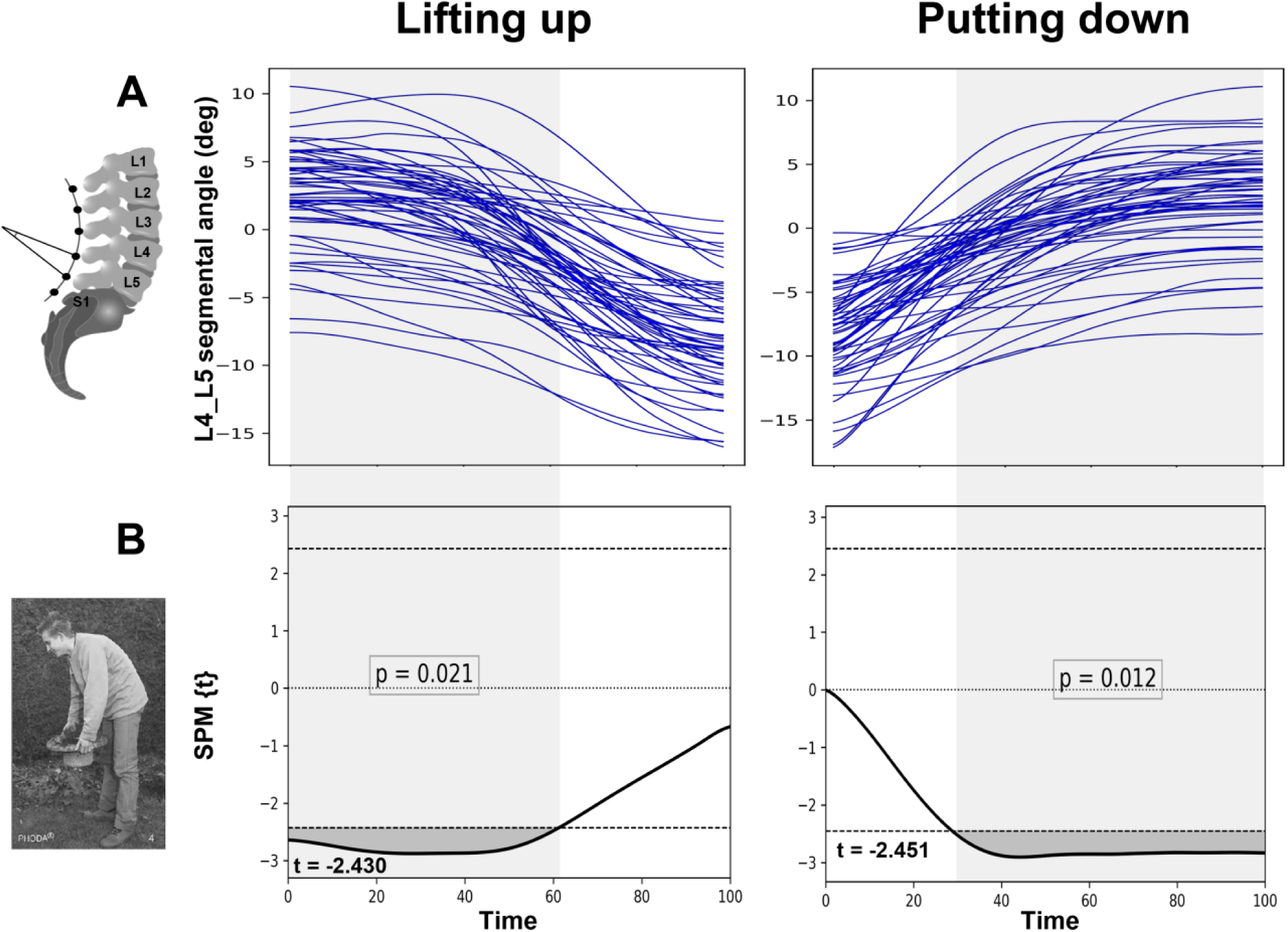
A = Individual (N = 57, time-normalized) continuous L4/L5 segmental angles during lifting-up (left) and putting-down (right) phases. X-axis: time normalized on 101 points (time window: 0 – 100%). B = t-statistics with supra-threshold clusters reflecting significant time-specific negative relationships between L4/L5 segmental angles during the two phases and the PHODA-lift (B) score, revealed by SPM1D multiple linear regression.

### 3.5. Relationships between PHODA-falling, PHODA-shoveling and continuous lumbar and thoracic angles during lifting

Using the PHODA-falling score as regressor of interest, a significant negative relationship to continuous lumbar angles was found during the lifting-up (time window: 0-77%, -0.484 < r > -0.319, p_FDR_ = 0.010) and putting-down phases (time window: 16-100%, -0.466 < r > -0.302, p_FDR_ = 0.005) (Figure 2A and 2C). Furthermore, the PHODA-falling score showed a significant negative relationship to the motion in almost all lumbar segments during both lifting phases (see Table 3). No significant relationships were found between thoracic angles and the PHODA-falling score, nor between the PHODA-shoveling score and continuous lumbar and thoracic angles (p_FDR_ > 0.05, see Table 1).

**Table 3:** Relationships between measures of pain-related fear and continuous segmental lumbar angles during lifting (uncorrected p-values)

| Lifting phase | Spinal region | Regressor | t-value | r-value | p-value |
| --- | --- | --- | --- | --- | --- |
| Lifting up | T11/L1 | PHODA-lift | 2.361 |  | 1.000 |
|  | L1/L2 |  | 2.410 |  | 0.324 |
|  | L2/L3 |  | 2.444 |  | 0.224 |
|  | L3/L4 |  | 2.463 |  | 0.364 |
|  | L4/L5 |  | 2.430 | -0.333 < r > -0.315 | **0.021** |
|  | L5/S1 |  | 2.451 |  | 0.281 |
|  | T11/L1 | PHODA-falling | 2.355 | -0.316 < r > -0.303 | **0.040** |
|  | L1/L2 |  | 2.408 | -0.342 < r > -0.312 | **0.025** |
|  | L2/L3 |  | 2.442 | -0.359 < r > -0.314 | **0.016** |
|  | L3/L4 |  | 2.462 | -0.424 < r > -0.318 | **0.016** |
|  | L4/L5 |  | 2.434 | -0.465 < r > -0.315 | **0.017** |
|  | L5/S1 |  | 2.455 | -0.334 < r > -0.316 | **0.013** |
| Putting down | T11/L1 | PHODA-lift | 2.381 |  | 0.644 |
|  | L1/L2 |  | 2.417 |  | 0.702 |
|  | L2/L3 |  | 2.459 |  | 0.657 |
|  | L3/L4 |  | 2.485 |  | 0.591 |
|  | L4/L5 |  | 2.451 | -0.354 < r > -0.305 | **0.012** |
|  | L5/S1 |  | 2.450 |  | 0.281 |
|  | T11/L1 | PHODA-falling | 2.377 |  | 0.889 |
|  | L1/L2 |  | 2.416 |  | 0.706 |
|  | L2/L3 |  | 2.460 | -0.316 < r > -0.306 | **0.014** |
|  | L3/L4 |  | 2.486 | -0.400 < r > 0.309 | **0.008** |
|  | L4/L5 |  | 2.459 | -0.475 < r > -0.299 | **0.009** |
|  | L5/S1 |  | 2.457 | -0.340 < r > -0.301 | **0.007** |

### 3.6. Effects of movement-specific pain-related fear on continuous lumbar angles

When including the TSK-G score as nuisance variable in the regression model, the observed negative relationships between the PHODA-lift score and the continuous lumbar angles remained statistically significant for both lifting phases (lifting-up: time window: 9-89%, -0.310 < r > -0.307, p_uncorr_ = 0.008; putting-down: time window: 15-60%, -0.315 < r > -0.305, p_uncorr_ = 0.027).

Similarly, the negative relationships between the PHODA-falling score and the continuous lumbar angles remained statistically significant for both lifting phases (lifting-up: time window: 0-76%, -0.491 < r > -0.317, p_uncorr_ = 0.010; putting-down: time window: 15-100%, -0.472 < r > -0.306, p_uncorr_ = 0.005).

### 3.7. Relationships between lumbar ROM during lifting and measures of pain-related fear

The lumbar ROM during lifting did not show a relationship with the TSK-G score (r = – 0.006, p = 0.965). Regarding the PHODA items, only the PHODA-falling score showed a statistically significant correlation with lumbar ROM (r = -0.380, p = 0.004). The results from the correlation analysis between lumbar ROM during lifting and the different PHODA items are found in Table 4.

## 4. Discussion

Here, we investigated whether pain-related fear affects lumbar spine motion in pain-free individuals to obtain information on potential pre-existing psychomotor interactions that might become relevant in a future LBP episode (e.g. stiffening the spine to protect the back). To this end, we performed analyses of continuous and discrete (ROM) sagittal plane spinal kinematics during a load lifting task, which is often perceived as a dangerous activity for the back [5,6,11] and correlated these data with self-reports of pain-related fear commonly used in research and clinical practice to assess different types of pain-related fear (general and movement-specific).

### The impact of pain-related fear on spinal motion in pain-free adults

Our results support the evolving evidence that fear avoidance beliefs and associated pain-related fear exist in the pain-free population. Further, we demonstrate their impact on spinal motion by showing distinct and time-specific relationships with lumbar motion during a lifting maneuver. In contrast, no effects of pain-related fear on thoracic motion were observed. The results also indicate different sensitivities of pain-related fear questionnaires in explaining variance of lumbar motion during lifting. General measures of pain-related fear, such as the TSK-G or the average PHODA score (PHODA-total), did not show an association with lumbar motion during lifting. In contrast, movement-specific pain-related fear, reflected by subjective ratings of potentially harmful movements during daily activities (PHODA-lift, PHODA-falling), demonstrated time-specific relationships with lumbar motion, even after accounting for linear effects of the TSK-G. This partially agrees with a recently reported association of pain-related fear with lumbar ROM during lifting in chronic LBP patients [34]. In line with the current study, that study observed a significant negative relationship between movement-specific pain-related fear, reflected by the PHODA-lift item, and lumbar motion during a lifting task, supporting the construct validity of the PHODA-lift item. However, we only observed the above-mentioned relationship in the time-sensitive SPM analysis, but not when using discrete lumbar ROM values. This discrepancy might be explained by a more subtle association between the PHODA-lift score and lumbar spine motion in pain-free individuals compared to chronic LBP patients, emphasizing the added value of time-sensitive analyses [43].

In the current study, only the PHODA item showing a person falling backwards on the grass demonstrated a significant association with the lumbar ROM during lifting. The PHODA-falling item is the only picture in the PHODA-SeV questionnaire that depicts an immediate and obvious risk of injury and might therefore have a stronger sensitivity for changes of the lumbar ROM. Furthermore, the PHODA-falling item was the only item that correlated with trait anxiety, indicating that this item might represent a different, general anxiety-related construct compared to the PHODA-lift item. In support of this, the detailed SPM1D analysis yielded segment-spanning relationships between the PHODA-falling scores and lumbar spine angles (Table 3) in both lifting phases, suggesting widespread non-specific effects on lumbar segmental motion.

### A protective movement strategy with potential negative consequences?

The observed alterations in lumbar spine motion in pain-free individuals with “round-back danger beliefs” point towards a change of the trunk movement strategy during lifting. This change is most likely achieved through altered neuromuscular activation/coordination, consistent with reports describing a protective response (i.e. tight control strategy), characterized by stiffening lumbar segments through antagonistic muscle activation leading to increased spinal stability [7,15,39,48,65]. Protective responses have been observed in pain-free individuals during anticipation of experimental back pain, characterized by reduced activation of deep trunk muscles and increased activation of superficial trunk muscles [39], similar to observations in patients with recurrent LBP [18]. These changes in the trunk movement strategy and their potential impact on LBP are gaining increasing attention [21,61]. A protective strategy might be beneficial in the short term by avoiding further pain or injury but is suggested to predispose individuals to back problems in the long term through reductions in lumbar spine fine motor control, increased loading of spinal tissues and muscle fatigue, promoting pro-nociceptive mechanisms and injury [20,37,39,49,59,61]. In particular, there is a trade-off between the risk of LBP or injury associated with spinal tissue overload and the risk of spinal instability [16]. Increased trunk stiffness, reflected by reduced lumbar flexion, is achieved through voluntary antagonistic muscle co-contraction [30] improving spinal stability during lifting but also strongly increasing spinal loading [15,16]. Increased spinal loading is known for initiating or accelerating spinal tissue degeneration [32,47,58].

Interestingly, the PHODA-lift score was specifically associated with altered motion in the L4/L5 region, which is where the majority of lumbar disc herniations or degenerative changes such as degenerative spondylolisthesis occur [31,53]. The L4/L5 motion segment must bear heavy loads, making it more susceptible for developing injury- or degeneration-related pain as compared to other lumbar segments [10,66]. Individuals scoring high on the PHODA-lift item seem to particularly stiffen the L4/L5 motion segment during lifting, indicating a segment-specific protective movement strategy. This is likely realized through antagonistic co-contraction of spinal muscles, which then leads to increased intervertebral loads at the L4/L5 level. Hence, those individuals might be at risk for facilitation of degenerative processes and/or for an LBP episode. Furthermore, the protective behavior for the L4/L5 segment seems to occur primarily in the first half of the lifting-up phase and in the second half of the putting-down phase, which may be explained by particularly large moment arms of the upper body and box masses and the respective high muscular responses during tasks involving large flexion angles.

### Pre-existing beliefs about lifting

Our results indicate that protective movement strategies can be driven by beliefs about the harmfulness of daily activities such as lifting with a flexed spine, in the absence of (experimental) pain. These beliefs, often held and communicated by health care professionals and manual handling advisors [42], likely originate from earlier *in vitro* studies investigating the effects of loads on cadaveric spines [1,4] and *in vivo* studies measuring intradiscal pressure [40,41], which led to the conclusion that lifting weights with a flexed spine yields a higher risk for disk injuries and LBP, compared to lifting with a neutral spine [40,41]. However, more recent studies do not support this notion. Dreischarf and colleagues (2016) reported only a 4% difference in load between the two different lifting techniques using an instrumented vertebral body replacement [13]. A recent systematic review concluded that the current advice to avoid lumbar flexion during lifting to prevent LBP is not justified [50].

## Limitations

There are some limitations of the current study that need to be mentioned. The measurement of lumbar angles using skin markers is strictly speaking a measurement of the external shape of the back in the lumbar region rather than an actual measurement of the angles between the respective vertebral bodies. Previous research showed that these angles differ by about 20° [67]. This limits the direct comparison of the lumbar angles reported in the current study with other studies; however, it does not affect the results of our regression analyses since all participants were measured identically. Furthermore, the accuracy of predicted curvature angles might have been affected by accumulating soft tissue in more extended positions of the lumbar spine. However, previous research showed that such inaccuracies occur mainly in lumbar extensions of more than 40° [52] and because most of the lumbar lordosis angles during the important phases in the current study were below 40°, we do not expect that the observed psychomotor relationships were due soft tissue-related inaccuracies.

## Conclusion

Lifting heavy loads under certain conditions (e.g. being distracted or fatigued) might indeed pose strong risks for triggering an acute LBP episode [55] and specific lifting techniques might be essential in certain work-related and everyday life situations. Nonetheless, we argue that the importance of lifting with a neutral spine in everyday activities has been greatly exaggerated and might even have detrimental effects. The current study demonstrated that beliefs underlying pain-related fear of lifting with a flexed spine do affect lumbar segmental motion in pain-free adults. The pre-existing altered spinal kinematics in individuals with round-back danger beliefs might indicate a protective trunk movement strategy that can be dysfunctional in the long term by initiating potential pro-nociceptive mechanisms or amplifying pain in an LBP episode.

Current results provide a basis for future kinematic and biomechanical analyses in LBP patients, focusing on time-specific psychomotor relationships that might be relevant for the maintenance of LBP and/or development of chronic LBP. The results also emphasize the need to raise more awareness of potential negative implications of erroneous beliefs regarding lifting techniques in the public and health sector.

## Data Availability

Data available upon request

## 5. Acknowledgments

This research was supported by the Swiss National Science Foundation (SNF, Bern, Switzerland). Movement analysis was performed with support of the Swiss Center for Clinical Movement Analysis, SCMA, Balgrist Campus AG, Zürich. We especially thank Marina Hitz, Linard Filli and Marc Bolliger from the SCMA for their support. Finally, we would like to thank Lukas Connolly for the valuable comments on this manuscript.

